# Analysis of Depression, Anxiety, ADHD, and Behavior Problems among 6-17-Year-Old U.S. Children: Evidence from the National Survey of Children’s Health

**DOI:** 10.1101/2025.11.25.25340981

**Authors:** Md Mohsin, Sultan Mahmud, Shaila Nazneen, Razia Mustani, Emre Umucu, Yok-Fong Paat, Thenral Mangadu

**Affiliations:** College of Health Sciences, The University of Texas at El Paso, 500 W University Ave, Texas 79968, USA; International Centre for Diarrheal Disease Research, Bangladesh (icddr,b), 68 Shaheed Tajuddin Ahmed Ave, Dhaka 1212, Bangladesh; Environmental Science and Engineering Program, The University of Texas at El Paso, 500 W University Ave, Texas 79968, USA

**Keywords:** Mental Health, Depression, Anxiety, ADHD, Behavior Problems, Children, NSCH

## Abstract

**Background:** Child mental health is a critical public health concern in the United States. This study examines the multifaceted determinants of child mental health, including demographic, socioeconomic, family, healthcare access, and chronic health condition information.

**Methods:** This study utilized data from the National Survey of Children’s Health (NSCH). Univariate, bivariate, and multivariable logistic regression models were fitted to identify the determinants of each outcome variable.

**Results:** We pooled data from two years (2020 & 2021) of NSCH and included a nationally representative sample of 6- to 17-year-old U.S. children (N = 60,809; male, 51.1%; female, 48.9%; 12-17-year-old, 50.9%; 6-11-year-old, 49.1%). There was an increase in the prevalence of depression, anxiety, and ADHD/ADD among 6-17-year-old U.S. children from 2020 to 2021, while the prevalence of behavioral problems decreased during the same period (the prevalences were 4.8%, 10.7%, 8.8%, and 11% respectively for the year of 2020 and 5.3%, 11%, 7.9% and 11.6% in 2021). Our findings reveal significant gender disparities, with girls experiencing higher rates of depression (AOR: 1.64; 95% CI: 1.34-2.01, P <0.001) and anxiety (AOR: 1.62; 95% CI: 1.43-1.84, P <0.001). Teens (12-17 years) were more susceptible to depression and anxiety than younger children (6-11 years). Ethnic and racial disparities were observed, with lower rates of these conditions among non-Hispanic Asian and Hispanic/Latino children. Neighborhood support was a protective factor against anxiety and behavioral problems. Physical activity for at least 60 minutes every day was significantly associated with lower odds of depression (AOR: 0.51; 95% CI: 0.37-0.71; P <0.001) and anxiety (AOR: 0.67; 95% CI: 0.54 - 0.83; P <0.001) as opposed to no physical activities. Adverse childhood experiences (ACEs) were consistently linked to a higher likelihood of mental health conditions.

**Conclusion:** This study underscores the complex interplay of factors influencing child mental health and offers actionable insights for policy and practice. Future research should explore causal relationships and longitudinal trajectories to guide effective interventions and policies.

## Introduction

Mental health problems in children are highly prevalent and a significant public health concern in the United States. The mental health of children has drawn increasing attention in recent years. This heightened attention is driven by the recognition that mental health issues during childhood can exert profound and lasting effects on cognitive, emotional, and social development, with potential consequences that reverberate into adulthood ^1–3^. Mental disorders in children are defined as significant deviations from how children usually behave, learn, or manage their emotions, which can be distressing and make it difficult to get through the day ^4^. Depression, anxiety, attention deficit hyperactivity disorder (ADHD), and behavior problems are the most common mental illnesses that can be identified in children and adolescents ^5^.

In 2010, 20% of U.S. children suffered from mental health issues ^6^. In 2016, 3.2% of U.S. children aged 3 to 17 experienced depression, 7.1% had anxiety, and 7.4% had behavior issues^7^. In addition, 9.4% of U.S. children aged 2 to 17 had been identified as having ADHD ^8^. In 2013-2019, among children aged 3-17 years, the prevalence of ADHD, anxiety, behavior problems, and depression was 9.8%, 9.4%, 8.9%, and 4.4%, respectively ^9^.

The COVID-19 pandemic might have significantly impacted children’s worsening mental health conditions due to school closures and the lack of outdoor activities. Study shows that in 2021, more than a third (37%) of high school students reported having poor mental health during the COVID-19 pandemic, and 44% said they had experienced chronic sadness or hopelessness during the previous year ^10^. According to a recent study using the National Survey of Children’s Health (NSCH) datasets, before the pandemic (2016–2019), there were apparent increases in anxiety and depression, with slight but statistically insignificant continuations of these trends in 2020. However, between 2019 and 2020, there was a considerable rise in conduct or behavior problems ^11^.

Despite widespread attention to reducing disparities, there are still gaps in the mental health status and treatment of youth from racial and ethnic minorities ^12^. Differences also exist in accessing mental health services due to socioeconomic factors. For example, compared to Latinx children, White youth were more than twice as likely to use psychotropic drugs. Disparities between Latinx-White and Black-White patients who filled prescriptions for psychiatric medications continued over time ^13^. White adolescents were estimated to have received more mental health care in a specialist setting in the previous year (17.2%) than Hispanic, Black, and Asian adolescents (13.2%, 11.8%, and 9.5%, respectively) ^14^.

Mental health illnesses are common, but they are not always obvious. Mental illnesses may go unnoticed, undetected, and untreated. According to reports, more than 60% of young people who are depressed do not receive medical care ^15^. These untreated illnesses can have severe and prolonged repercussions on a person’s health. Long-term untreated mental illnesses may increase the risk of diabetes, heart disease, and stroke ^16^. They may also cause suicidal ideation and increase the risk of suicide, the 10^th^ leading cause of death in the U.S. ^15^.

Despite progress in understanding and addressing mental health issues in children, there is a clear need for an updated and comprehensive analysis, particularly due to the COVID-19 pandemic. This manuscript fills a significant gap in existing literature by thoroughly examining the mental health challenges faced by 6 to 17-year-old children in the United States. It relies on robust data from the National Survey of Children’s Health (NSCH), conducted periodically by the CDC. The NSCH’s extensive nationwide dataset, which includes demographic, socioeconomic, family, healthcare access, and chronic health condition information, enables a detailed exploration of the various risk and protective factors influencing children’s mental health outcomes. The theory of ecological systems ^17^, the theory of social disorganization ^18^, and the idea of social determinants of health ^19^ all formed the basis for the conceptual framework of this study.

## Methods

### Data Source and Participants

The present study used data from the National Survey of Children’s Health (NSCH), a nationally representative survey conducted annually by the U.S. Census Bureau. The NSCH employs a two-phase data collection methodology, including a preliminary household screener questionnaire and a detailed topical questionnaire completed by the parents or primary caregivers of a randomly selected child from each household. The survey is available in both online and paper formats and covers various topics, including children’s physical and mental health, access to healthcare, demographics, parental socioeconomic status, and neighborhood characteristics. The survey is financed and overseen by the Maternal and Child Health Bureau of the Health Resources and Services Administration ^20^.

To enhance participant recruitment, the NSCH implements various strategies. Firstly, a letter is sent to selected families inviting them to complete the survey online. If there is no response, they receive a paper survey after two reminder letters have been sent. Additionally, a phone line is established to assist participants who may require support completing the study or have any queries. These tactics aim to increase participation in the survey ^20^. Further information on the methodology employed for each year’s survey can be accessed here: ^21^.

We analyzed the 2020 and 2021 rounds of the NSCH datasets in this study, which are publicly available. We excluded children under six years old from the analyses. We used complex survey design weights and parameters in the analyses to make the estimates generalizable to all U.S. children aged 6-17 years.

### Outcome Variables

In this study, we examined four dichotomous (yes/no) outcome variables from independent survey questions related to whether the child has been diagnosed by a healthcare professional with depression, anxiety, ADD/ADHD, and behavior problems. These variables are crucial indicators of a child’s overall mental health and represent the most frequently diagnosed mental disorders among children ^5^. Past research and national estimates have indicated that these four conditions often coexist and are commonly identified concurrently in an individual ^20,22,23^.

### Covariates

According to the literature, various factors can impact children’s mental health at the child, family, neighborhood, and society levels. In the analysis, we considered several factors relating to children, adults, and families. We included covariates in this study based on previous studies on the same topic and datasets. We also had the covariates in the analyses if they seemed relevant to our outcome variables in the emerging scenarios due to the COVID-19 pandemic.

### Child-Related Covariates

Child-related covariates for this study are sex, age, race/ethnicity, ACE status, the number of times moved to a new address since born, birth weight, currently covered by health insurance, needs treatment for emotional, developmental, and behavioral problems, physical activity of at least 60 mins last week, Autism/ASD currently, etc.

Based on the National Center on Birth Defects and Developmental Disabilities’ classification of age groups at the Centers for Disease Control and Prevention ^24^, we recorded children’s ages into two categories: middle childhood (6–11 years old), and teens/teenagers (12–17 years old). We stratified our analyses by the age group of the children because the risk for mental health varies with age ^20,25^. We generated a composite variable for adverse childhood experiences based on the 10 ACE types that NSCH covers and have been used in earlier research ^20^. We categorized children as having no ACEs, having 1-2 ACEs, and having 3 or more ACEs. Previous studies followed this categorization for reporting ACEs ^26,27^.

### Familial and Other Covariates

Familial and other covariates include household size; the highest level of education among reported adults in the household; family poverty ratio; family structure; mental health problems among adults; neighborhood support level; anyone who smokes inside the home, etc.

### Statistical Analysis

Descriptive analysis was performed for each variable to assess the distribution of the variables. Logistic regression models with a generalized linear model (GLM) were fitted to identify the determinants of each of the outcomes. Only the significant variables in the unadjusted models were used in the final models. The 5% significance level was considered when estimating the odds ratios (OR) and confidence intervals (CI). These analyses were performed by applying a complex survey design of the NSCH in Stata v. 16.0 software (Stata Corporation LLC, College Station, TX) utilizing the survey subpopulation statistics (svy and sub pop commands).

### Ethical Considerations

This study analyzed publicly available secondary data. No identifiable data was used in this study. Details of the survey, data, and the study were shared with the Institutional Review Board of the University of Texas at El Paso (UTEP), and it was approved.

## Results

Table 1 presents the sociodemographic characteristics of the study population. According to the data, there were slightly more male children (51.1%) than female children (48.9%). Most children were in their teens (12-17 years old, 50.9%) as opposed to middle childhood (6-11 years old, 49.1%). Non-Hispanic white children represented the largest racial/ethnic group (49.8%), followed by Hispanic/Latino children (26.2%). Most children lived in households with five or more members (45.3%), and more than half had two biological/adoptive parents who were currently married (57.2%). The highest level of education among reported adults varied, with the majority having a college degree or higher (49.0%). Over 90% of children were covered by health insurance.

**Table 1:**
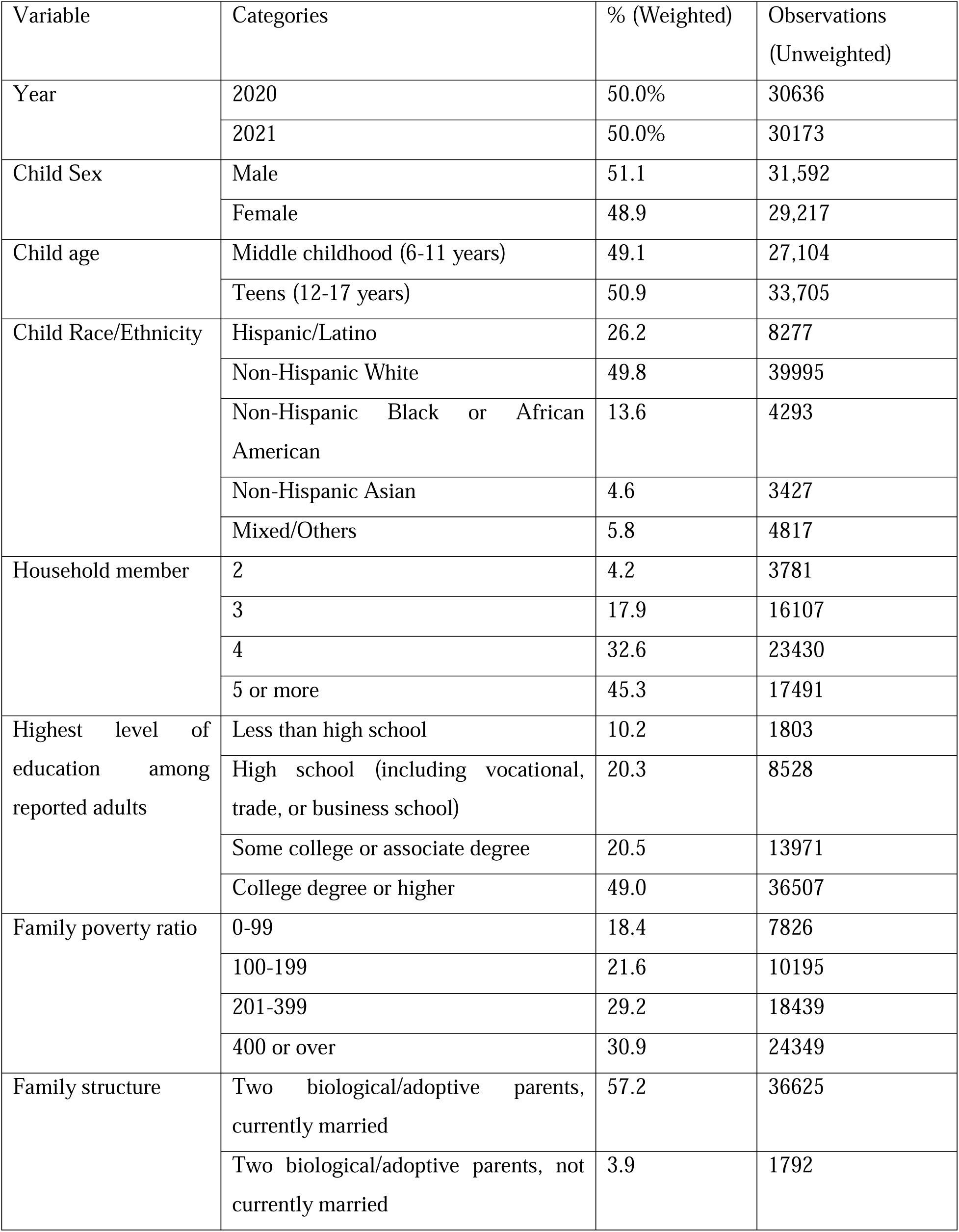
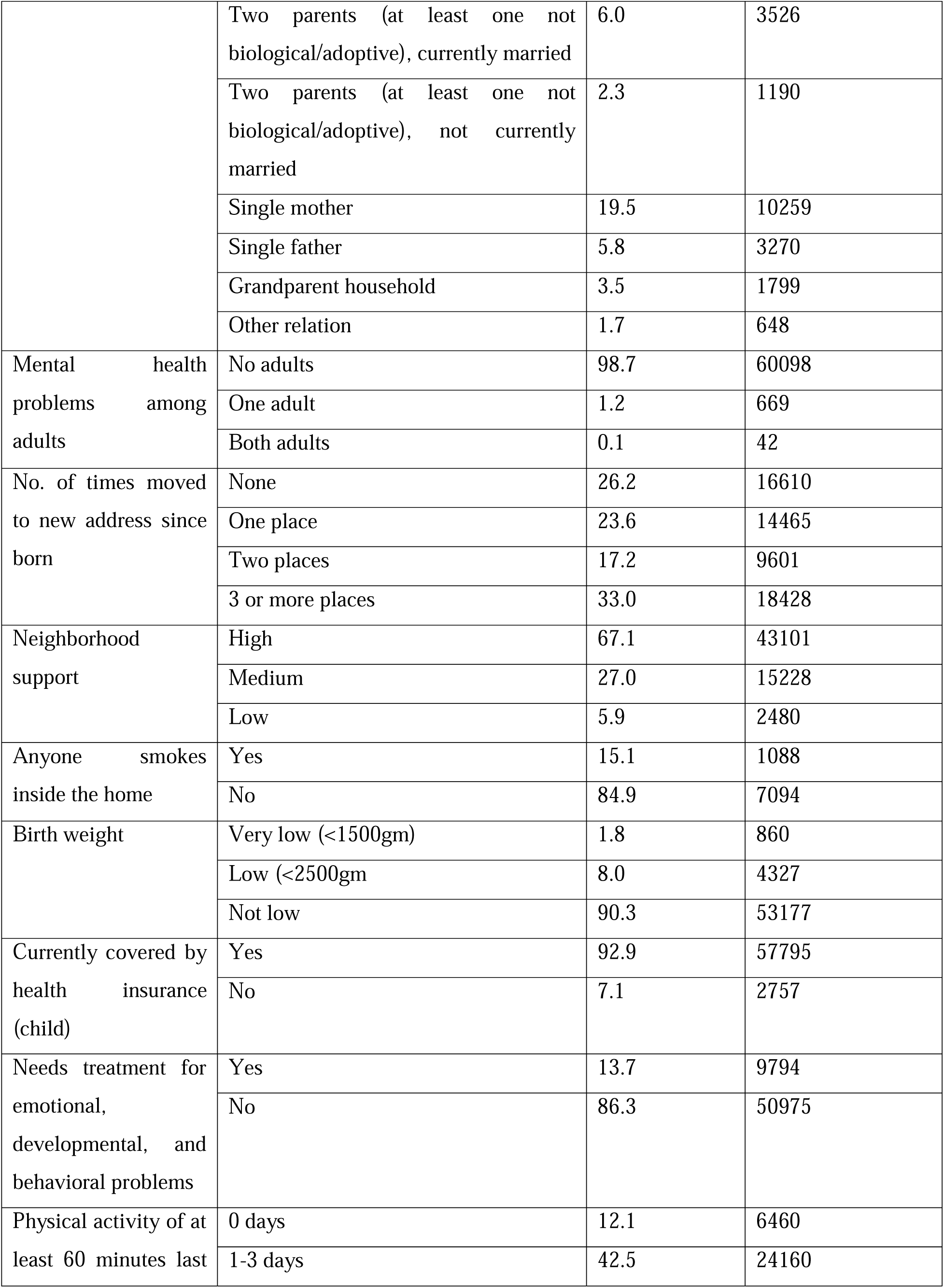
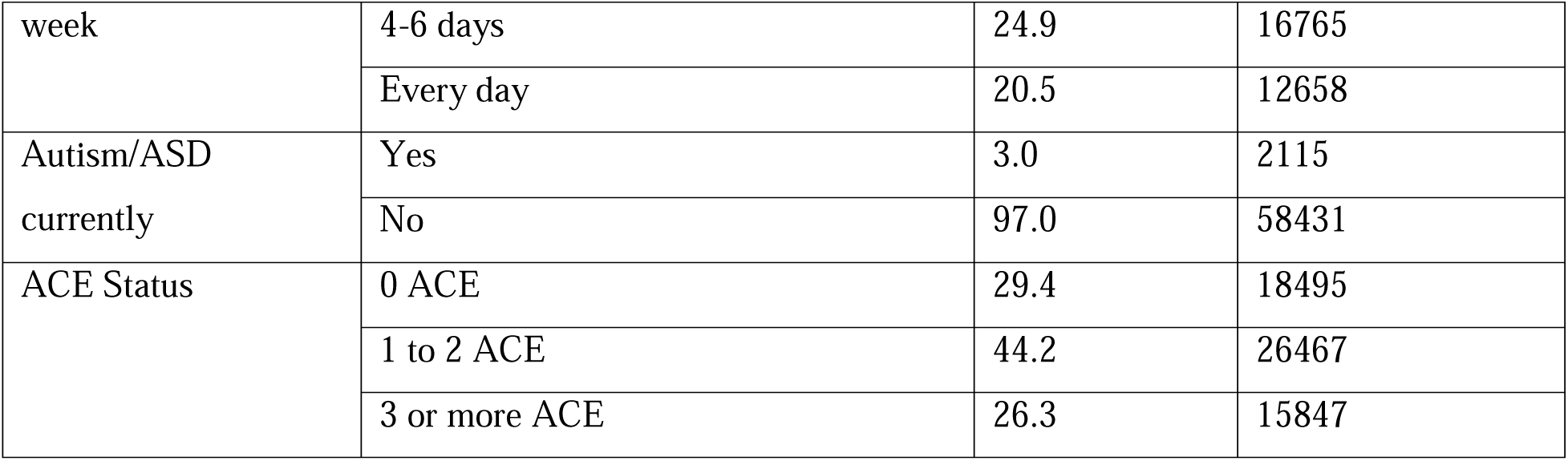
Sociodemographic characteristics of 6-17-year-old U.S. children, pooled estimates of NSCH 2020-2021, N=60,809.

The data also revealed that a small percentage of adults had mental health problems in the household (1.3%), and only 13.7% needed treatment for emotional, developmental, and behavioral issues. Additionally, 67.1% of children had high neighborhood support. Only 15.1% of children lived in households where someone smoked inside the home. Regarding physical activity, 42.5% of children engaged in at least 60 minutes for 1-3 days in the past week. Lastly, 29.4% of children had 0 ACE (Adverse Childhood Experience), 44.2% had 1 to 2 ACEs, and 26.3% had three or more ACEs.

Figure 1 shows an increase in the prevalence of depression, anxiety, and ADHD/ADD among 6-17-year-old U.S. children from 2020 to 2021, while the prevalence of behavioral problems decreased during the same period. Figure 2 depicts the 2020 and 2021 prevalence of these conditions in two age groups: middle childhood (ages 6-11) and teens (ages 12-17). In middle childhood, depression decreased from 1.9% (2020) to 1.5% (2021), while anxiety remained stable at 7.2% (2020) and 7.1% (2021). Behavioral problems were higher than depression and anxiety, at 9.9% (2020) and 7.9% (2021), while ADHD/ADD remained at 9.9% in both years. Among teens, depression increased from 7.7% (2020) to 9.1% (2021), and anxiety remained relatively steady at 14.1% (2020) and 14.8% (2021). In 2021, behavioral problems remained stable as in 2020, and ADHD/ADD increased from 12.6% (2020) to 13.6% (2021).

**Figure 1:**
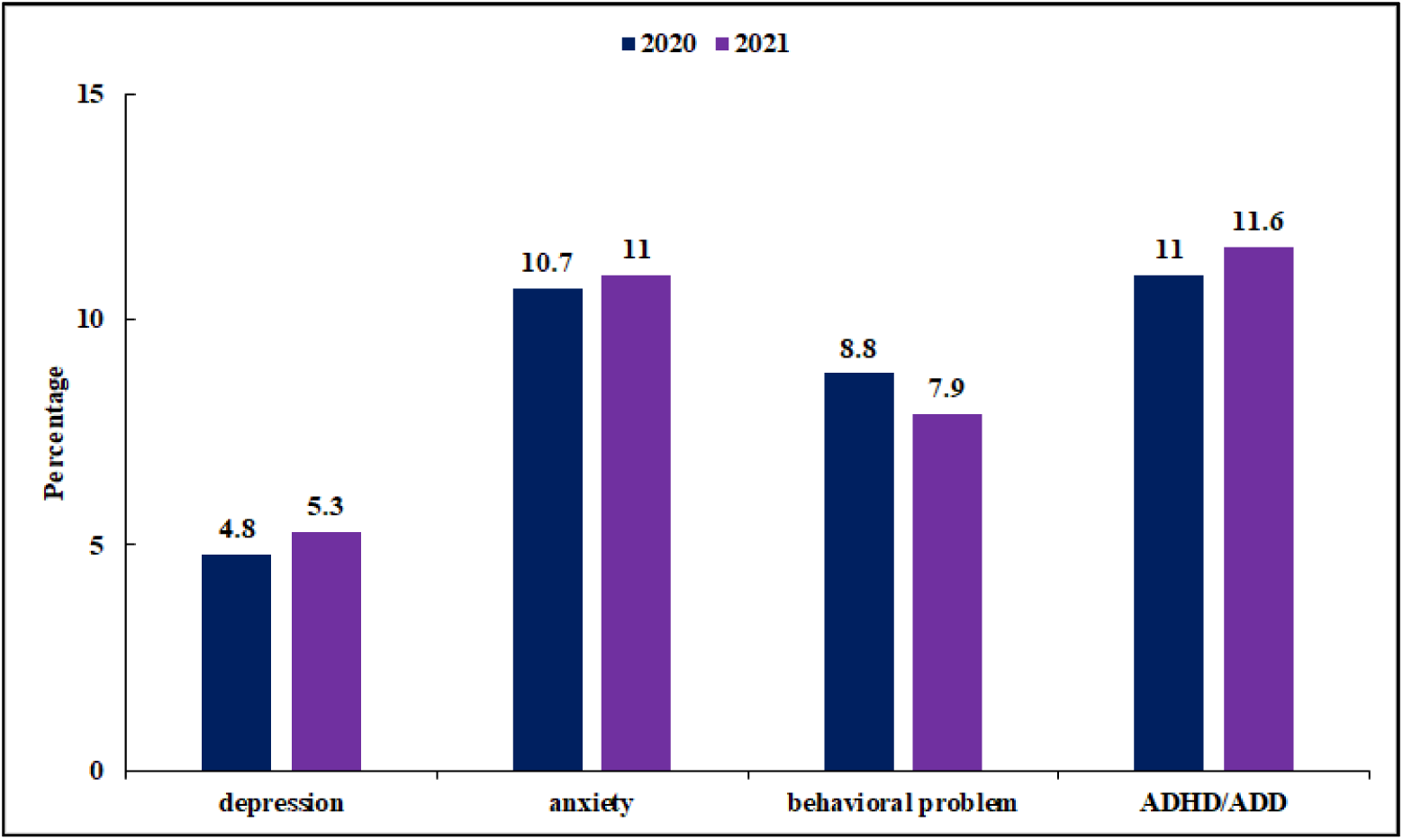
Prevalence of Depression, Anxiety, Behavior Problems, and ADHD/ADD among 6-17-Year-Old U.S. Children (NSCH 2020-2021, N=60,809) by Year.

**Figure 2:**
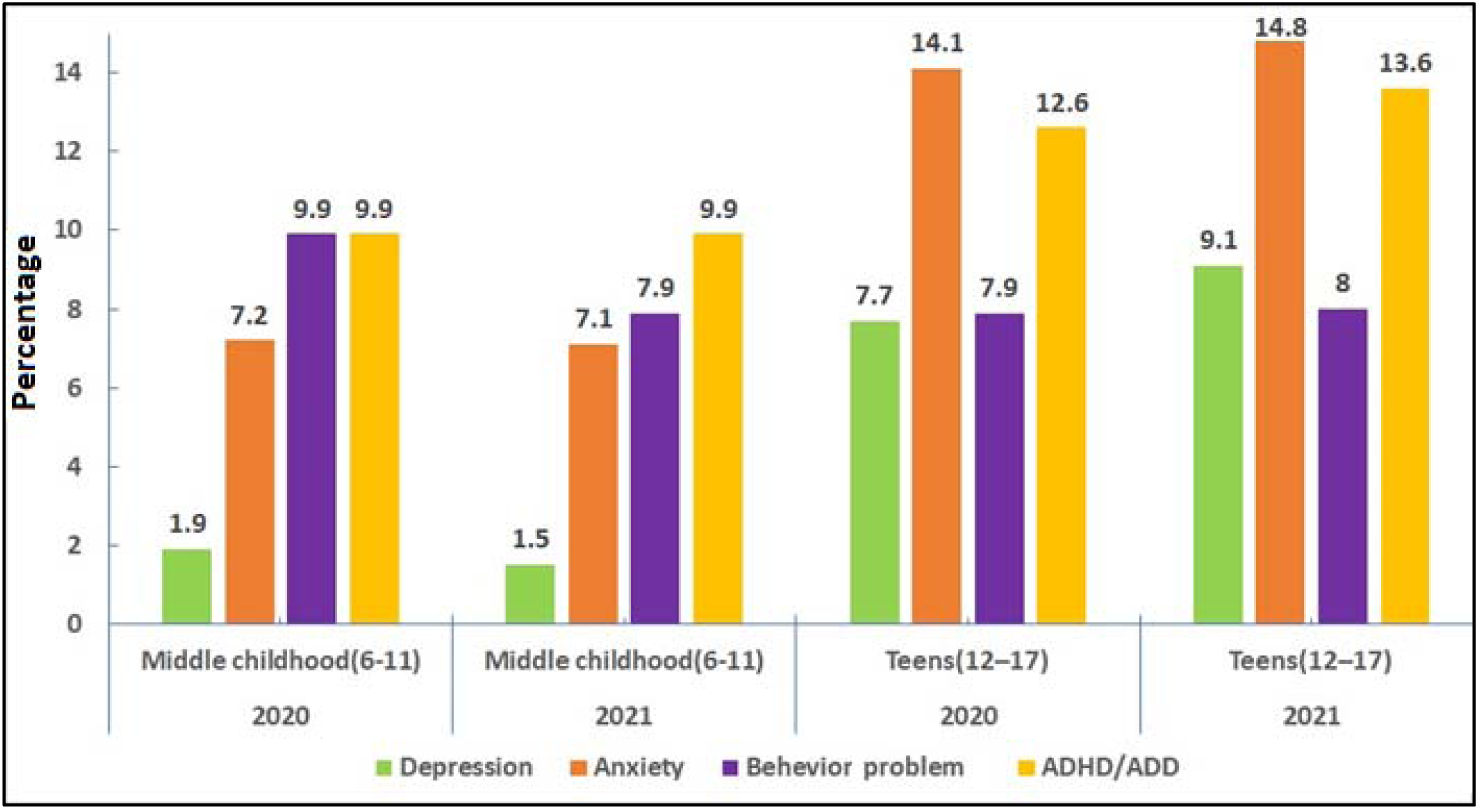
Prevalence of Depression, Anxiety, Behavior Problems, and ADHD/ADD among 6-17-Year-Old U.S. Children (NSCH 2020-2021, N=60,809) by Age group and Year.

Table 2 displays the unadjusted and adjusted associates of depression children aged 12-17 years had 5.58 (95% CI: 4.37, 7.13) times higher odds of developing depression compared to children aged 6-11 years. Female children were 1.64 (95% CI: 1.34, 2.01) times more likely to suffer from depression than male children. Non-Hispanic Asians had a 66% (AOR=0.44, 95%CI: 0.28, 0.69) reduced likelihood of experiencing depression than non-Hispanic Whites. Children with single fathers had a 41% (AOR: 0.59, 95% CI: 0.37, 0.96) lower probability of being depressed compared to those with two biological/adoptive parents who are currently married. Children who participated in physical activity for at least 60 minutes for 1-3 days, 4-6 days, and every day in the last week had respectively 30% (AOR: 0.71, 95% CI: 0.56, 0.90), 50% (AOR: 0.50, 95% CI: 0.39, 0.68), and 49% (AOR: 0.51, 95% CI: 0.37, 0.71) lower odds of developing depression compared to children who did not engage in physical activity in the last week. The odds of having depression among children with 1 to 2 Adverse Childhood Experiences (ACEs) were 1.80 (95% CI: 1.13, 2.87) times higher, while the odds of depression among children with three or more ACEs were 4.19 (95% CI: 2.33, 7.52) times higher than those with no ACEs.

**Table 2:**
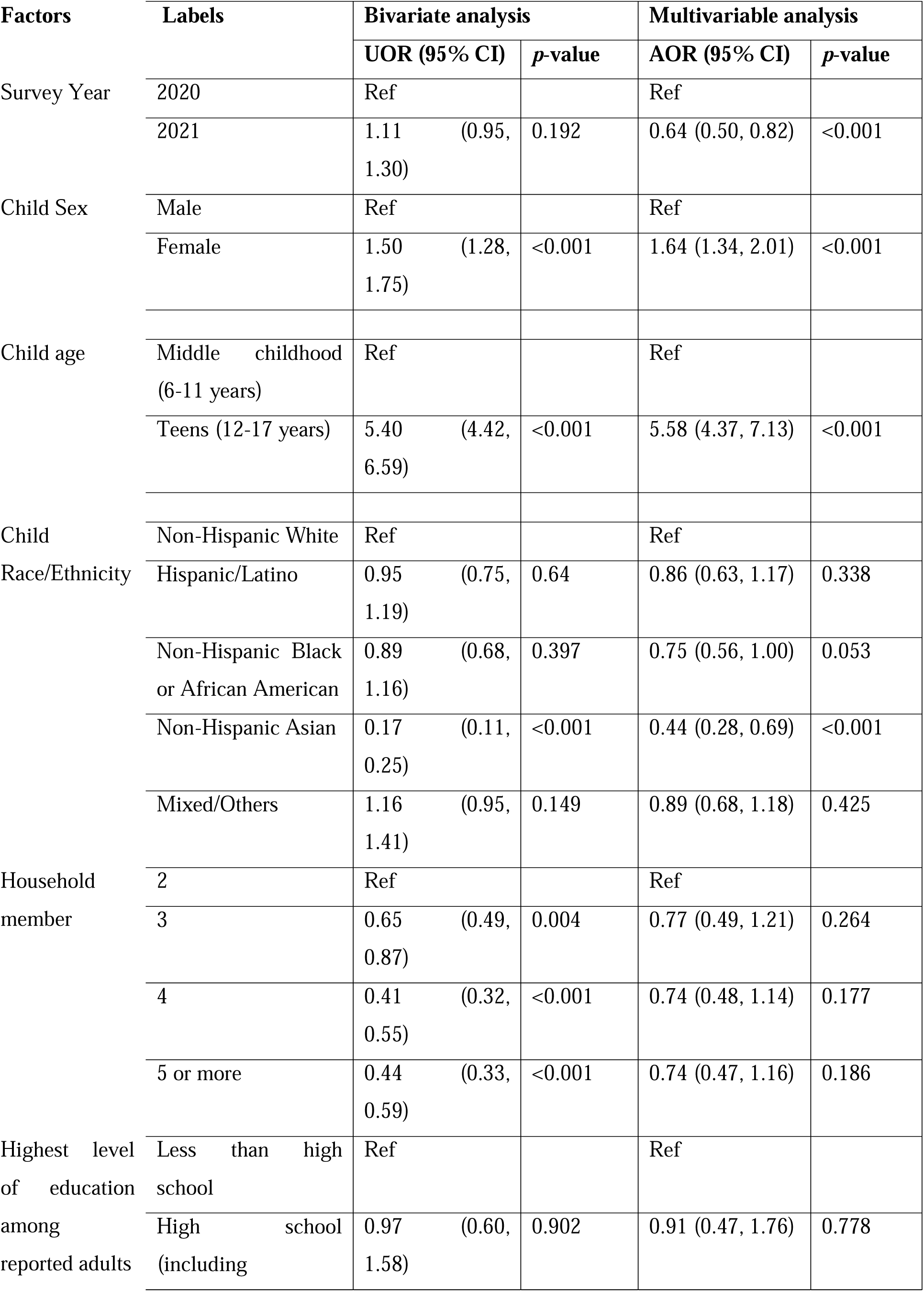
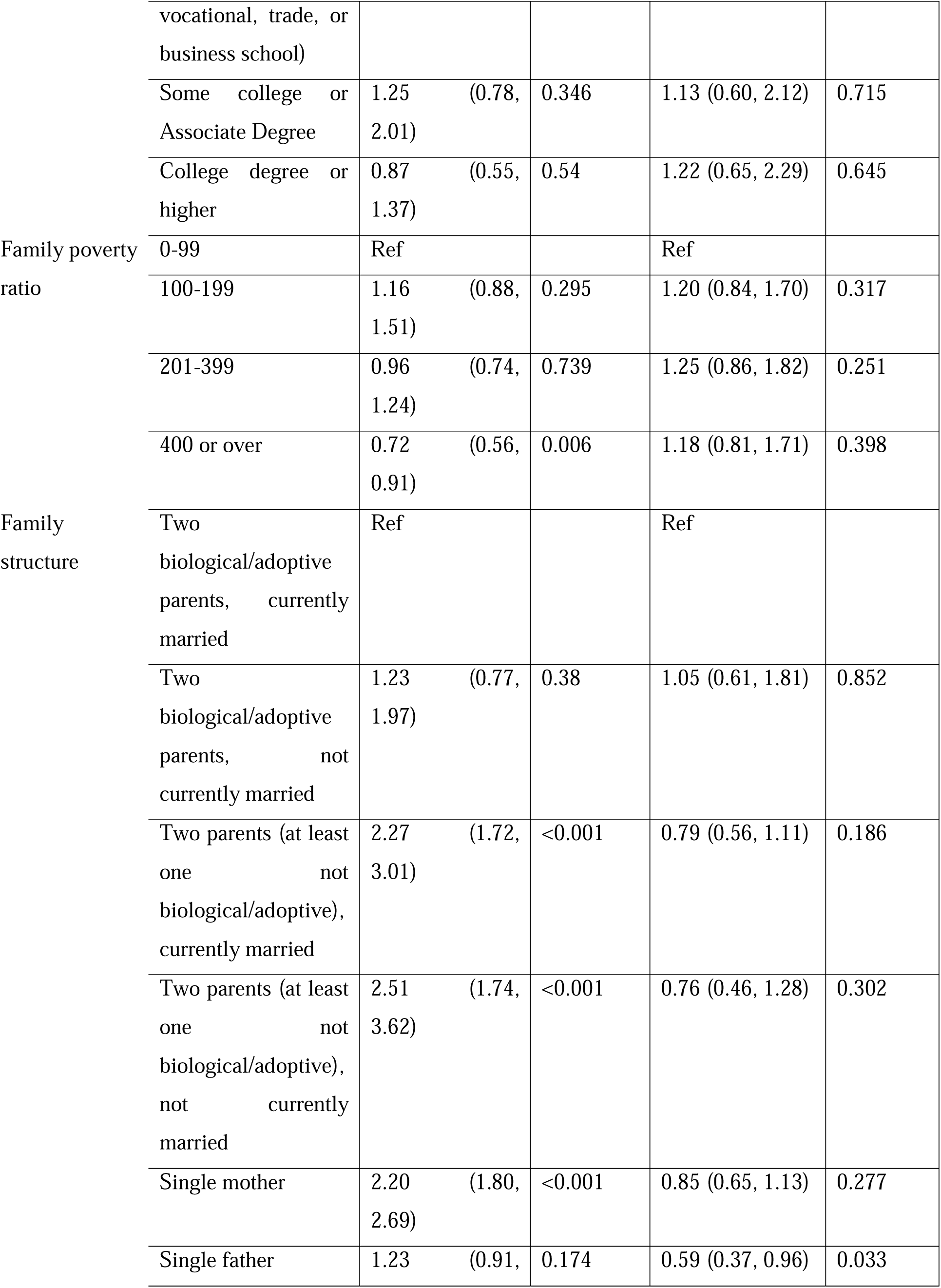
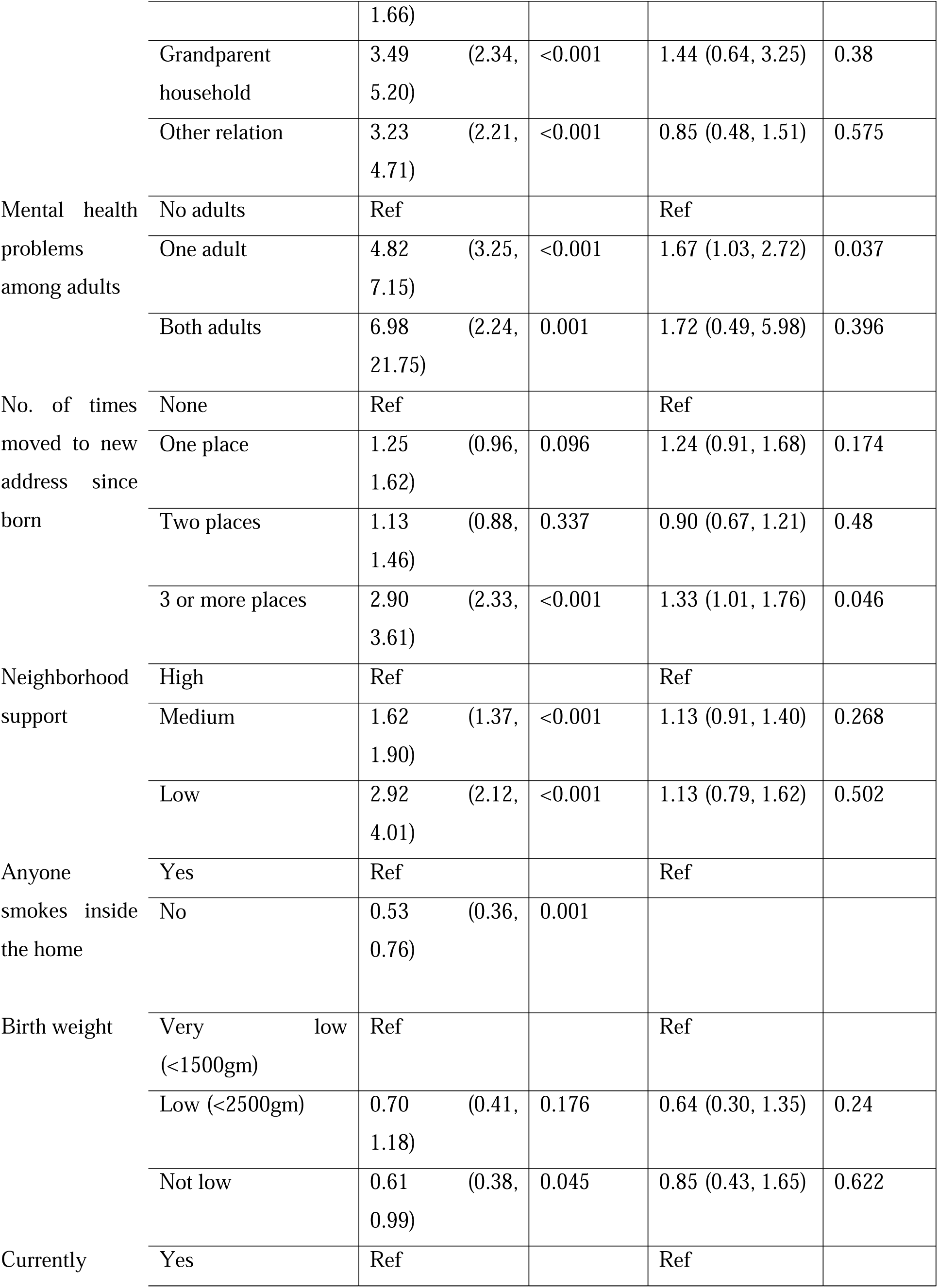
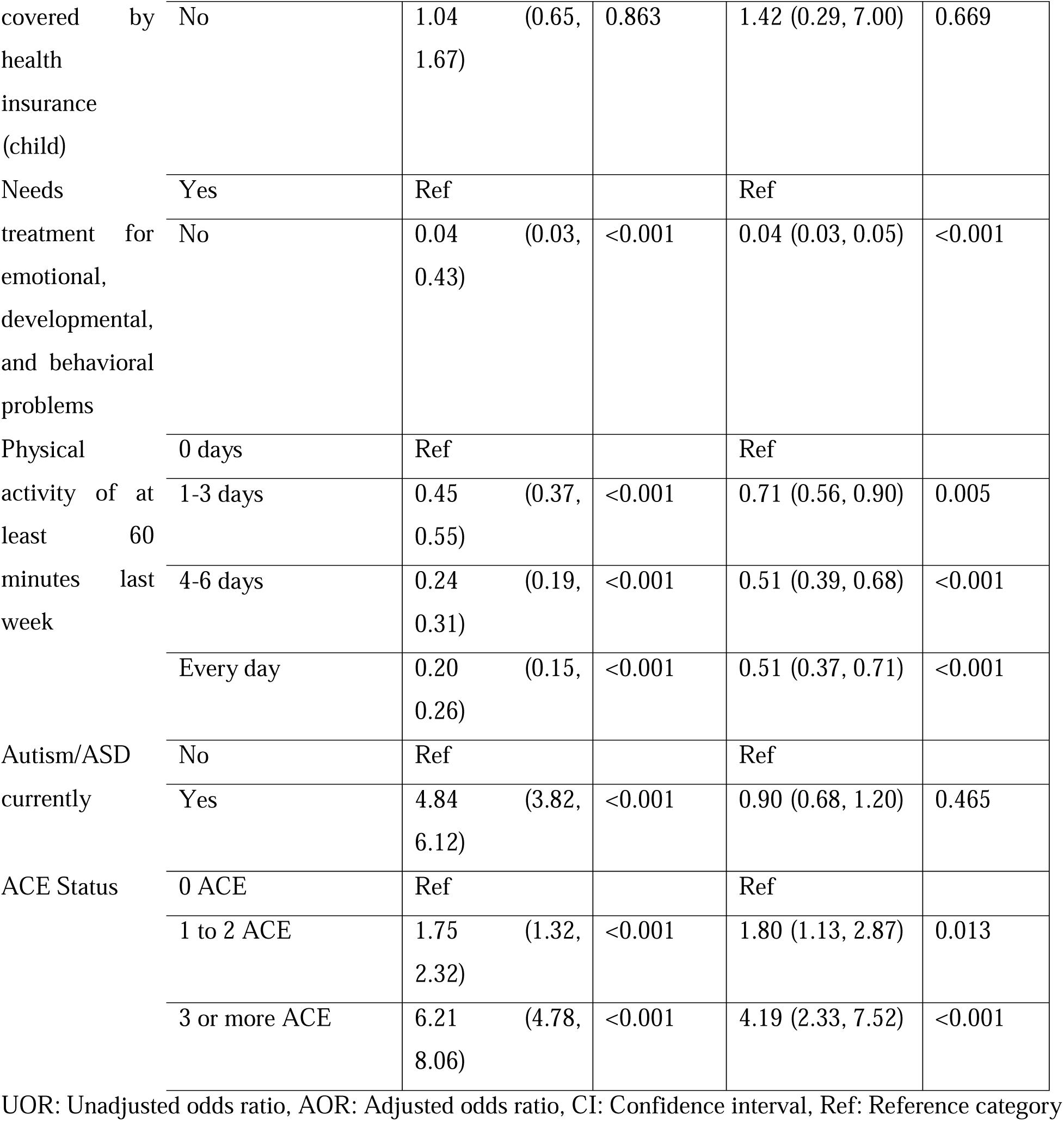
Unadjusted and adjusted associates of depression among 6–17-year-old U.S. children, pooled estimates of NSCH 2020-2021, N=60,809.

Table 3 shows the unadjusted and adjusted correlates of anxiety among U.S. children aged 6-17. The results indicate that children between the ages of 12-17 years old had a significantly higher odds ratio of anxiety (AOR = 2.16, 95% CI: 1.90,2.46) compared to those in the middle childhood age group (6-11 years old). Furthermore, female children were 1.62 (95% CI: 1.43, 1.84) times more likely to suffer from anxiety than male children. Additionally, Hispanic/Latino children had 41% (AOR=0.59, 95% CI: 0.49, 0.73) lower odds of experiencing anxiety compared to non-Hispanic Whites, while Non-Hispanic Black or African American children had 59% (AOR=0.41, 95% CI: 0.33, 0.51) lower odds and Non-Hispanic Asians had a 68% (AOR=0.44, 95% CI: 0.28, 0.69) lower odds of anxiety.

**Table 3:**
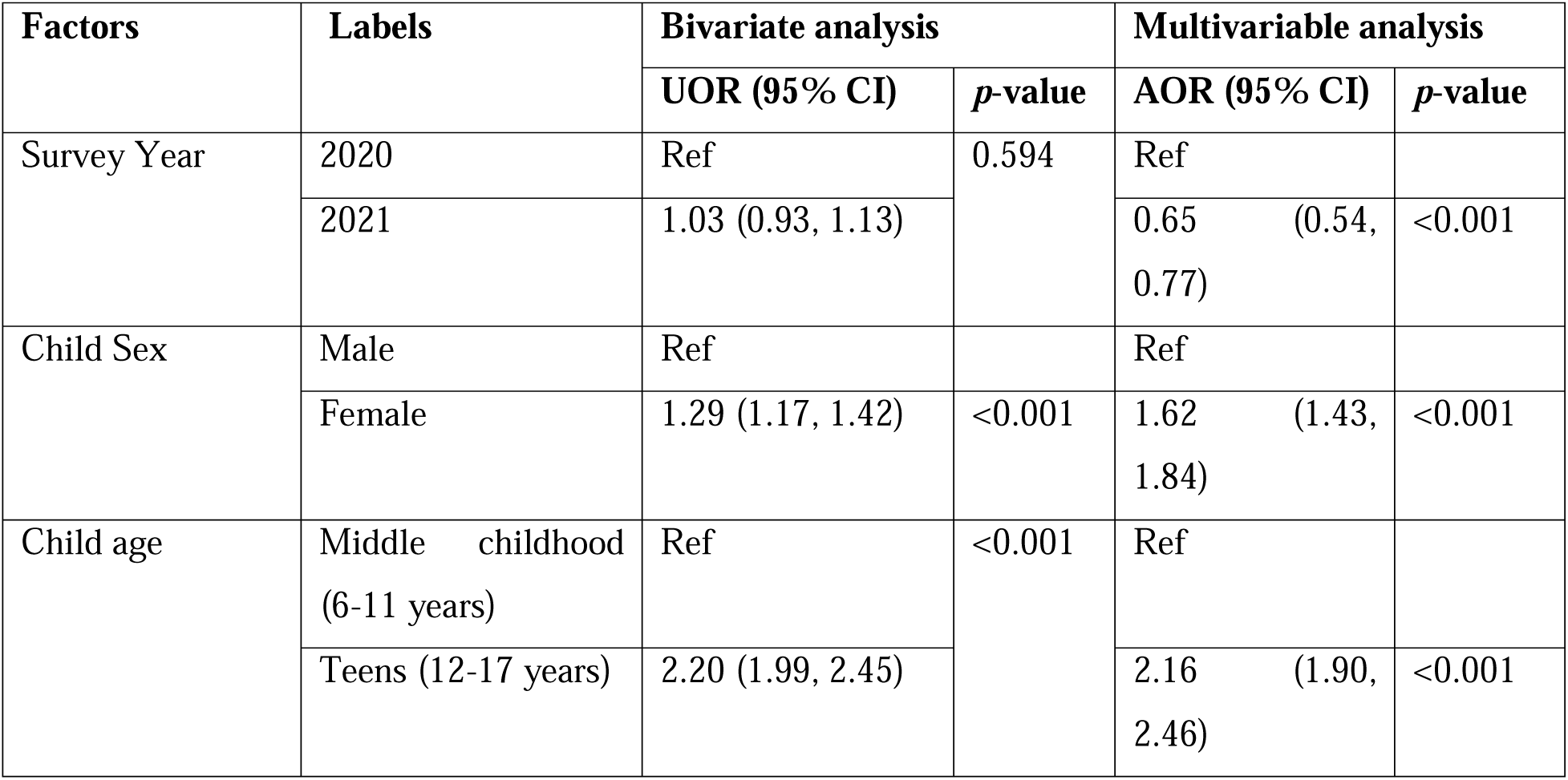
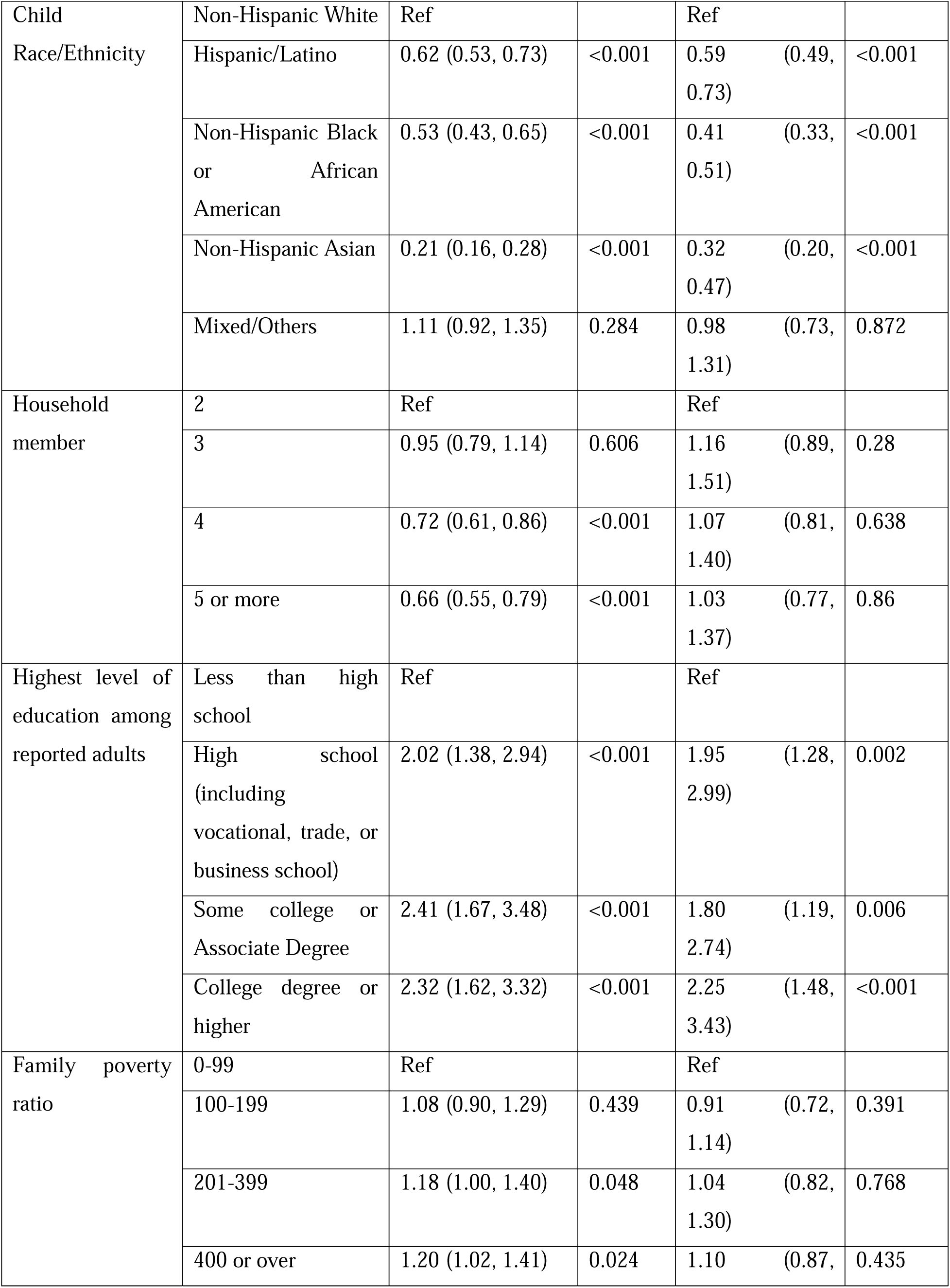
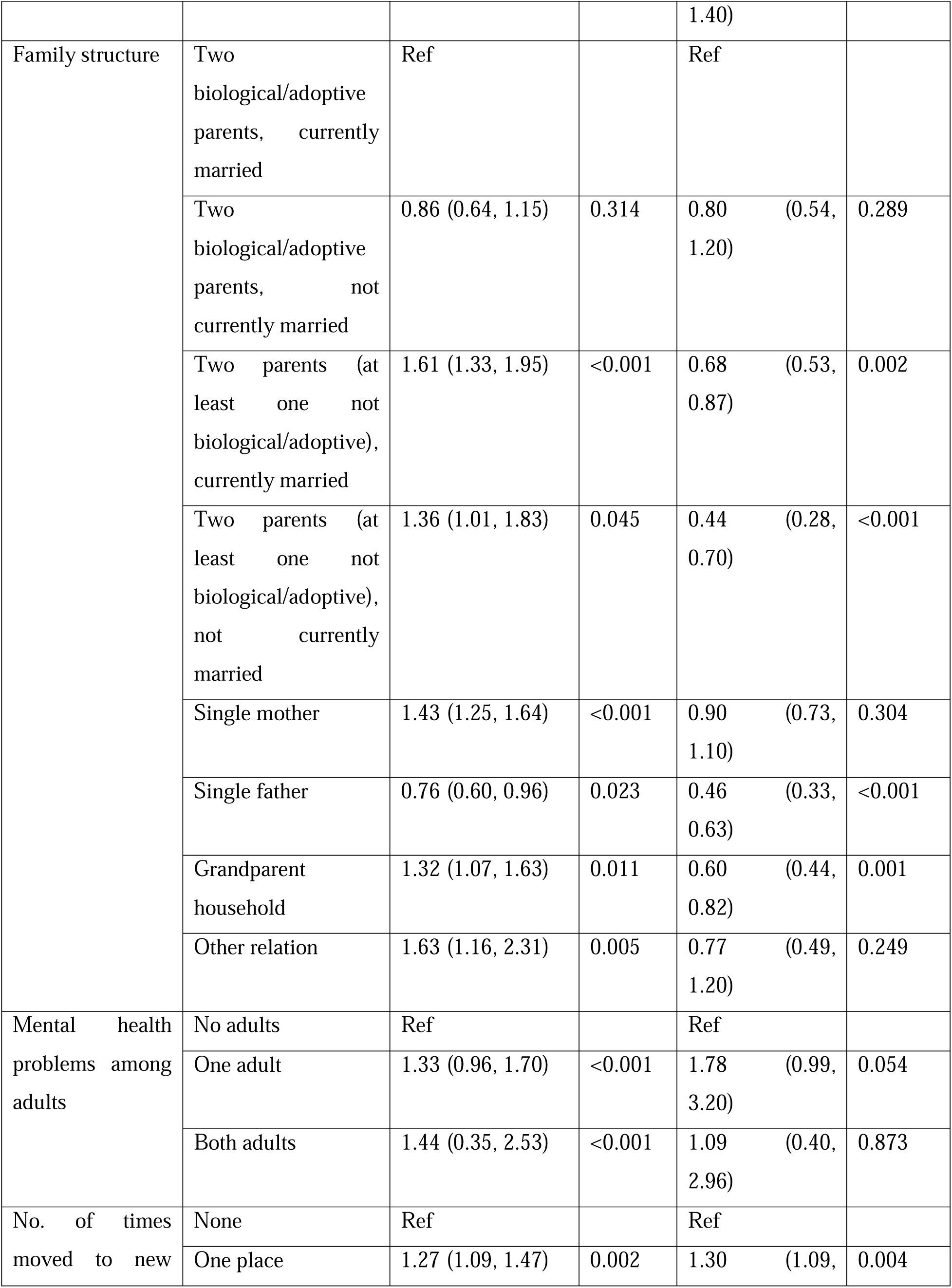
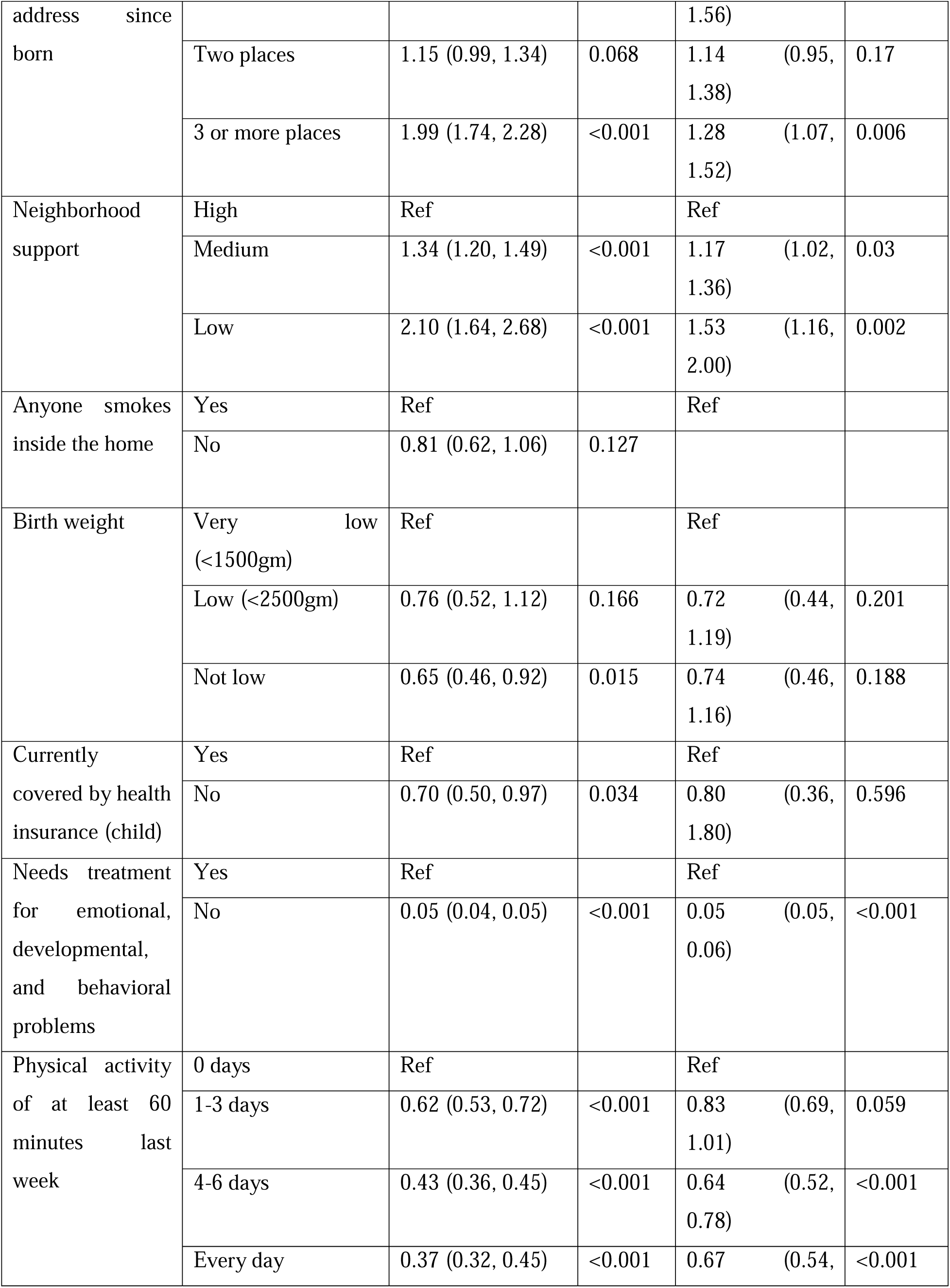
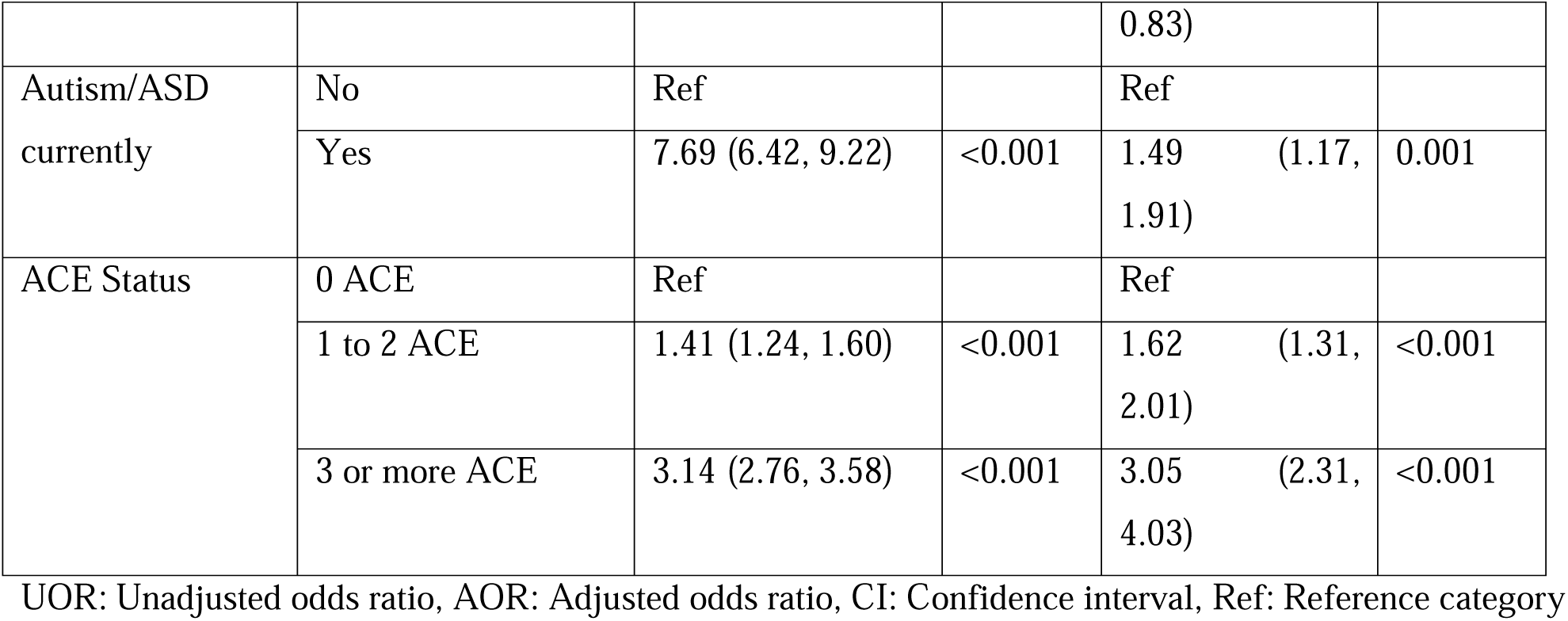
Unadjusted and adjusted associates of anxiety among 6–17-year-old U.S. children, pooled estimates of NSCH 2020-2021, N=60,809.

Moreover, the study found that children who live with single fathers or in grandparent households had a lower probability of experiencing anxiety than those with two biological/adoptive parents currently married. Specifically, the odds were 54% lower (AOR: 0.46, 95% CI: 0.33, 0.63) for children with single fathers and 40% lower (AOR: 0.60, 95% CI: 0.44, 0.82) for those living in grandparent households.

The study also found that children with low or medium neighborhood support were respectively 1.17 (95% CI: 1.01, 1.36) and 1.53 (95% CI: 1.162)) times more likely to suffer from anxiety compared to children who had higher support. In contrast, children who engaged in physical activity for at least 60 minutes for 4-6 days and every day in the last week had respectively 37% (AOR: 0.63, 95% CI: 0.52, 0.78) and 33% (AOR: 0.67, 95% CI: 0.54, 0.83) lower odds of developing anxiety compared to those who did not engage in physical activity in the last week.

Furthermore, children diagnosed with Autism/ASD at the time of data collection had a 1.53 (95% CI: 1.16, 2.00) times higher chance of suffering from anxiety. Finally, the odds of having depression were found to be 1.62 (95% CI: 1.31, 2.01) times higher among children with 1 to 2 Adverse Childhood Experiences (ACEs). In comparison, the odds of anxiety were 3.05 (95% CI: 2.31, 4.03) times higher among children with three or more ACEs than those without ACEs.

Table 4 portrays the unadjusted and adjusted correlation of behavioral problems among children aged 6-17 years in the U.S. The children between 12-17 years had a 37% (AOR=0.63, 95% CI: 0.53,0.75) lower likelihood of having behavioral problems than the children between 6-11 years old. Female kids had a 55% (AOR= 0.45, 95% CI: 0.37,0.54) lower likelihood of having behavioral problems than male kids. Moreover, Hispanic/Latino children had a 34% (AOR=0.0.66, 95% CI: 0.51,.87) lower odds of experiencing behavioral problems compared to non-Hispanic Whites, while Non-Hispanic Asians had 60% (AOR=0.40, 95% CI: 0.28, 0.59) lower odds of behavioral problems.

**Table 4:** Unadjusted and adjusted associates of Behavior problems among 6–17-year-old U.S. children, pooled estimates of NSCH 2020-2021, N=60,809.

| Factors | Labels | Bivariate analysis |  | Multivariable analysis |  |
| --- | --- | --- | --- | --- | --- |
|  |  | UOR (95% CI) | *p*-value | AOR (95% CI) | *p*-value |
| Survey Year | 2020 | Ref |  | Ref |  |
|  | 2021 | 0.89 (0.78, 1.01) | 0.064 | 0.72 (0.57, 0.90) | 0.004 |
| Child Sex | Male | Ref |  | Ref |  |
|  | Female | 0.46 (0.40, 0.53) | <0.001 | 0.45 (0.37, 0.54) | <0.001 |
| Child age | Middle childhood (6-11 years) | Ref |  | Ref |  |
|  | Teens (12-17 years) | 0.88 (0.78, 1.00) | 0.05 | 0.63 (0.53, 0.75) | <0.001 |
| Child Race/Ethnicity | Non-Hispanic White | Ref |  | Ref |  |
|  | Hispanic/Latino | 0.82 (0.67, 0.99) | 0.041 | 0.66 (0.51, 0.87) | 0.003 |
|  | Non-Hispanic Black or African American | 1.33 (1.11, 1.59) | 0.002 | 0.95 (0.75, 1.22) | 0.701 |
|  | Non-Hispanic Asian | 0.24 (0.18, 0.32) | <0.001 | 0.40 (0.28, 0.59) | <0.001 |
|  | Mixed/Others | 1.41 (1.11, 1.79) | 0.005 | 1.12 (0.84, 1.49) | 0.455 |
| Household member | 2 | Ref |  | Ref |  |
|  | 3 | 0.81 (0.63, 1.05) | 0.11 | 1.26 (0.93, 1.71) | 0.135 |
|  | 4 | 0.60 (0.47, 0.76) | <0.001 | 1.29 (0.93, 1.79) | 0.127 |
|  | 5 or more | 0.65 (0.51, 0.84) | 0.001 | 1.42 (1.01, 1.99) | 0.044 |
| Highest level of education among reported adults | Less than high school | Ref |  | Ref |  |
|  | High school (including vocational, trade, or business school) | 1.48 (1.03, 2.12) | 0.034 | 1.15 (0.70, 1.87) | 0.589 |
|  | Some college or Associate Degree | 1.69 (1.19, 2.42) | 0.004 | 1.24 (0.77, 1.99) | 0.382 |
|  | College degree or higher | 1.03 (0.73, 1.45) | 0.867 | 1.21 (0.75, 1.93) | 0.438 |
| Family poverty ratio | 0-99 | Ref |  | Ref |  |
|  | 100-199 | 0.92 (0.75, 1.13) | 0.445 | 0.95 (0.72, 1.24) | 0.684 |
|  | 201-399 | 0.69 (0.57, 0.83) | <0.001 | 0.94 (0.71, 1.24) | 0.655 |
|  | 400 or over | 0.52 (0.43, 0.63) | <0.001 | 0.98 (0.73, 1.30) | 0.862 |
| Family structure | Two biological/adoptive parents, currently married | Ref |  | Ref |  |
|  | Two biological/adoptive parents, not currently married | 1.37 (0.99, 1.89) | 0.058 | 1.04 (0.73, 1.49) | 0.833 |
|  | Two parents (at least one not biological/adoptive), currently married | 2.09 (1.68, 2.60) | <0.001 | 1.23 (0.91, 1.66) | 0.171 |
|  | Two parents (at least one not biological/adoptive), not currently married | 3.26 (2.15, 4.93) | <0.001 | 1.65 (0.87, 3.14) | 0.129 |
|  | Single mother | 2.36 (2.00, 2.79) | <0.001 | 1.26 (0.95, 1.66) | 0.104 |
|  | Single father | 1.21 (0.92, 1.59) | 0.17 | 0.96 (0.67, 1.37) | 0.816 |
|  | Grandparent household | 3.04 (2.41, 3.82) | <0.001 | 1.35 (0.93, 1.96) | 0.116 |
|  | Other relation | 4.40 (2.96, 6.52) | <0.001 | 2.37 (1.19, 4.72) | 0.014 |
| Mental health problems among adults | No adults | Ref |  | Ref |  |
|  | One adult | 1.45 (0.99, 1.90) | <0.001 | 1.73 (0.79, 3.79) | 0.171 |
|  | Both adults | 1.17 (0.04, 2.30) | 0.042 | 1.21 (0.38, 3.86) | 0.753 |
| No. of times moved to new address since born | None | Ref |  | Ref |  |
|  | One place | 1.17 (0.96, 1.43) | 0.12 | 1.18 (0.93, 1.49) | 0.17 |
|  | Two places | 1.33 (1.08, 1.63) | 0.008 | 1.13 (0.87, 1.47) | 0.361 |
|  | 3 or more places | 2.45 (2.06, 2.92) | <0.001 | 1.36 (1.09, 1.68) | 0.005 |
| Neighborhood support | High | Ref |  | Ref |  |
|  | Medium | 1.79 (1.56, 2.05) | <0.001 | 1.54 (1.27, 1.86) | <0.001 |
|  | Low | 3.17 (2.48, 4.06) | <0.001 | 1.89 (1.39, 2.56) | <0.001 |
| Anyone smokes inside the home | Yes | Ref |  | Ref |  |
|  | No | 0.55 (0.40, 0.75) | <0.001 |  |  |
| Birth weight | Very low (<1500gm) | Ref |  | Ref |  |
|  | Low (<2500gm) | 0.79 (0.52, 1.19) | 0.26 | 1.20 (0.69, 2.07) | 0.523 |
|  | Not low | 0.63 (0.44, 0.90) | 0.011 | 1.39 (0.84, 2.30) | 0.2 |
| Currently covered by health insurance (child) | Yes | Ref |  | Ref |  |
|  | No | 0.94 (0.64, 1.38) | 0.738 | 3.23 (1.31, 7.98) | <0.011 |
| Needs treatment for emotional, developmental, and behavioral problems | Yes | Ref |  | Ref |  |
|  | No | 0.03 (0.02, 0.29) | <0.001 | 0.03 (0.03, 0.04) | <0.001 |
| Physical activity of at least 60 minutes last week | 0 days | Ref |  | Ref |  |
|  | 1-3 days | 0.59 (0.48, 0.71) | <0.001 | 0.80 (0.62, 1.03) | 0.088 |
|  | 4-6 days | 0.48 (0.39, 0.60) | <0.001 | 0.91 (0.64, 1.27) | 0.566 |
|  | Every day | 0.62 (0.50, 0.77) | <0.001 | 0.95 (0.72, 1.26) | 0.735 |
| Autism/ASD currently | No | Ref |  | Ref |  |
|  | Yes | 17.23 (14.29, 20.77) | <0.001 | 2.3113 (1.77, 3.02) | <0.001 |
| ACE Status | 0 ACE | Ref |  | Ref |  |
|  | 1 to 2 ACE | 1.41 (1.12, 1.67) | <0.001 | 1.23 (0.94, 1.57) | 0.129 |
|  | 3 or more ACE | 3.37 (2.87, 3.97) | <0.001 | 1.52 (1.06, 2.17) | 0.024 |

UOR: Unadjusted odds ratio, AOR: Adjusted odds ratio, CI: Confidence interval, Ref: Reference category

Moreover, the findings stated that children with low or medium neighborhood support were respectively 1,885 (95% CI: 1.39, 2.56) and 1.54 (95% CI: 1.27,1.86) times more likely to experience behavioral problems than those with higher support. Furthermore, children who had moved to three or more new addresses since birth were 1.36 (95% CI: 1.09, 1.68) times more likely to experience behavioral problems than those without a history of changing addresses. Additionally, children diagnosed with Autism/ASD at the time of data collection had a 2.31 (95% CI: 1.77, 3.02) times higher chance of experiencing behavioral problems. Finally, the odds of experiencing behavioral problems were 1.52 (95% CI: 1.06, 2.17) times higher among children with three or more Adverse Childhood Experiences (ACEs) than those without ACEs.

However, the findings of the GLM model were unsuccessful in establishing a significant association between family structure and behavioral problems among children. Additionally, at least 60 minutes of physical activity remained insignificant in influencing behavioral issues among children.

Table 5 shows the findings of the generalized linear model to highlight the association between numerous factors and the history of ADHD/ADD among children in the U.S. age range of 6-17 years. The children between 12-17 years had 1.255 (95% CI: 1.11, 1.42) times higher odds of experiencing ADHD/ADD than those aged 6-11. Female children had a 55% (AOR= 0.45, 95% CI: 0.39,0.54) lower probability of having ADHD/ADD than male children. Moreover, Hispanic/Latino children had a 39% (AOR=0.62, 95% CI: 0.50, .76) reduced likelihood of experiencing ADHD/ADD as opposed to non-Hispanic Whites, while non-Hispanic black or African American children had 29% (AOR= 0.71, 95% CI: 0.58,0.86) lower odds and Non-Hispanic Asians had 79% (AOR=0.22, 95% CI: 0.15,0.31) lower odds of having ADHD/ADD.

**Table 5:**
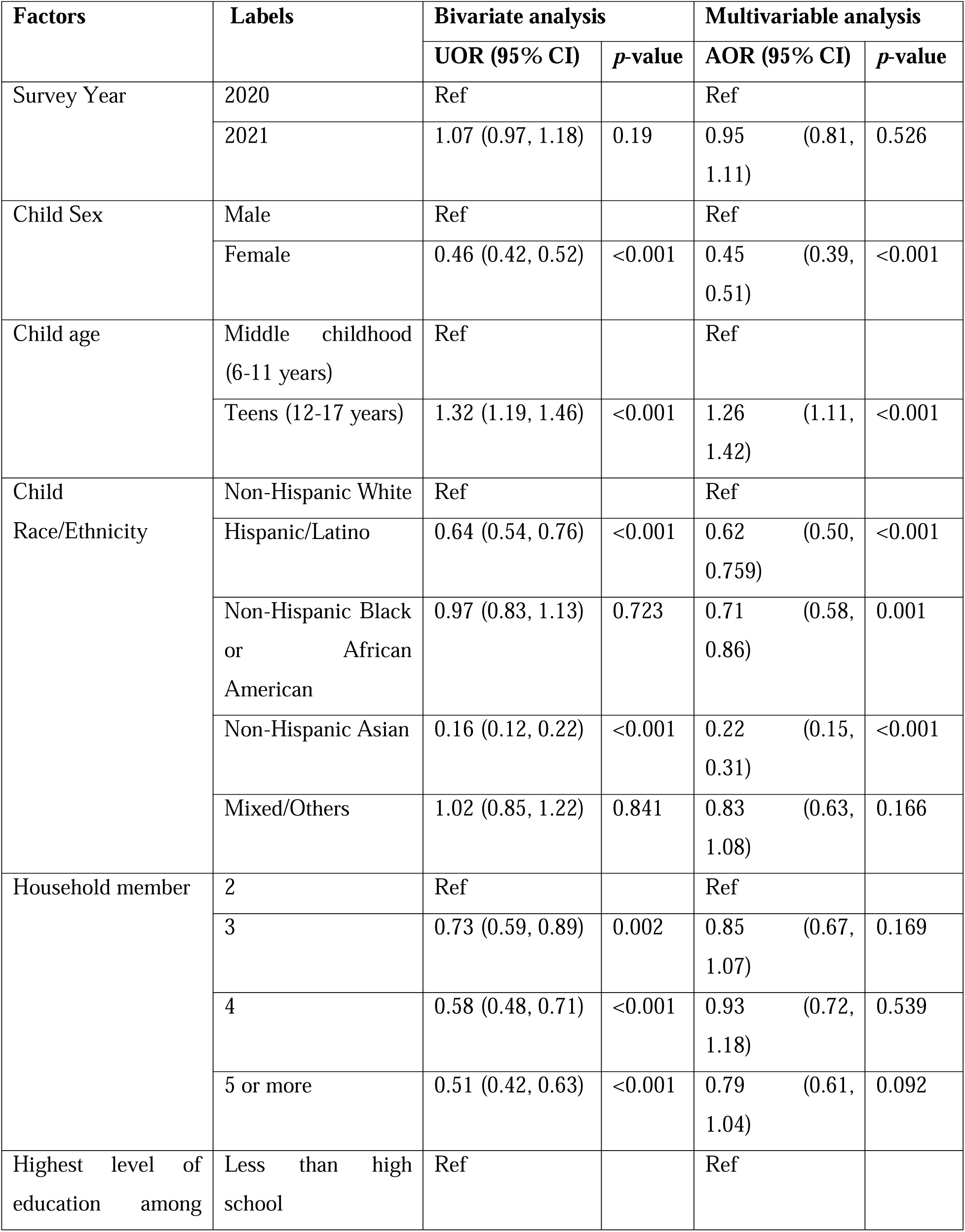
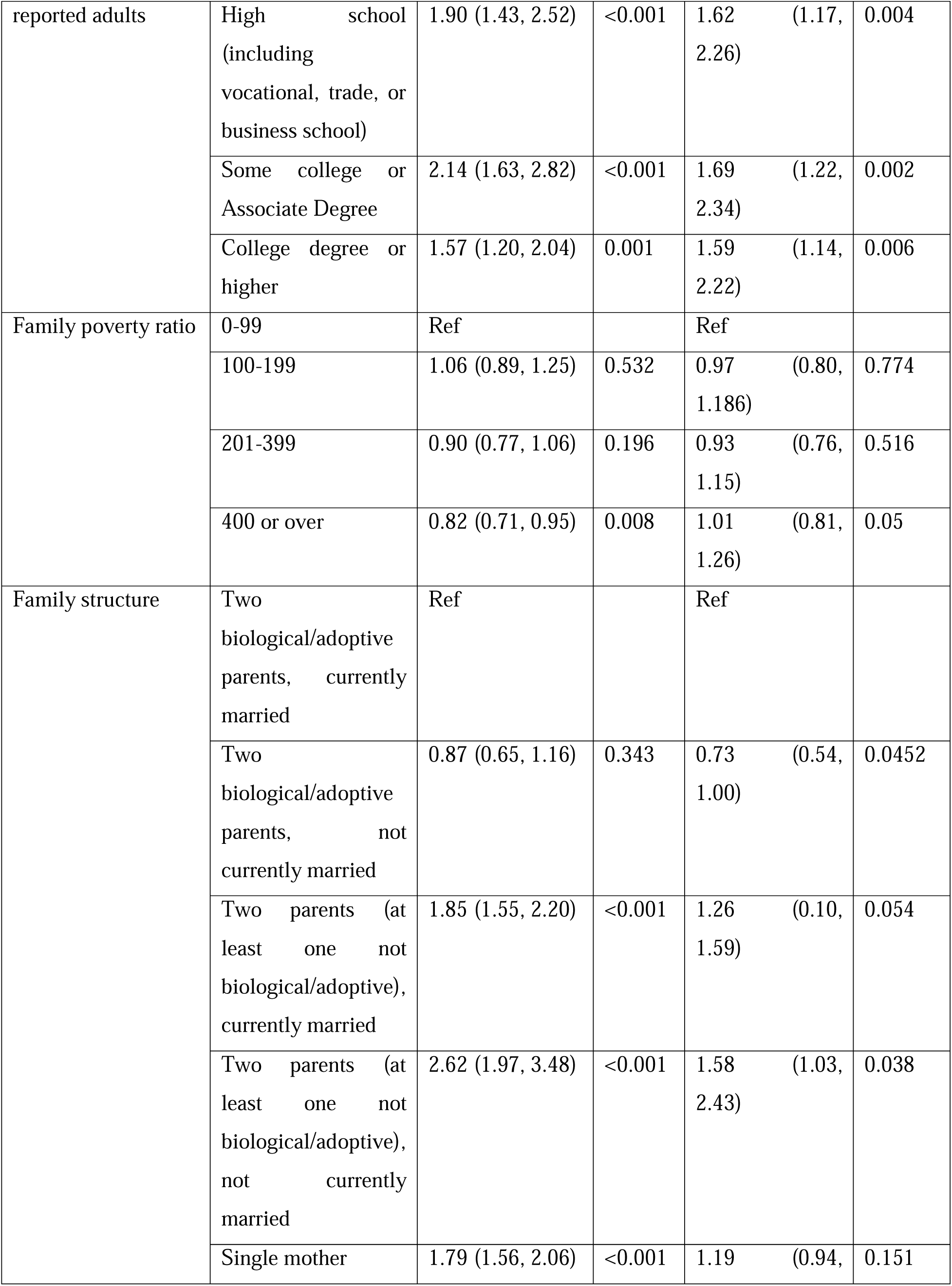
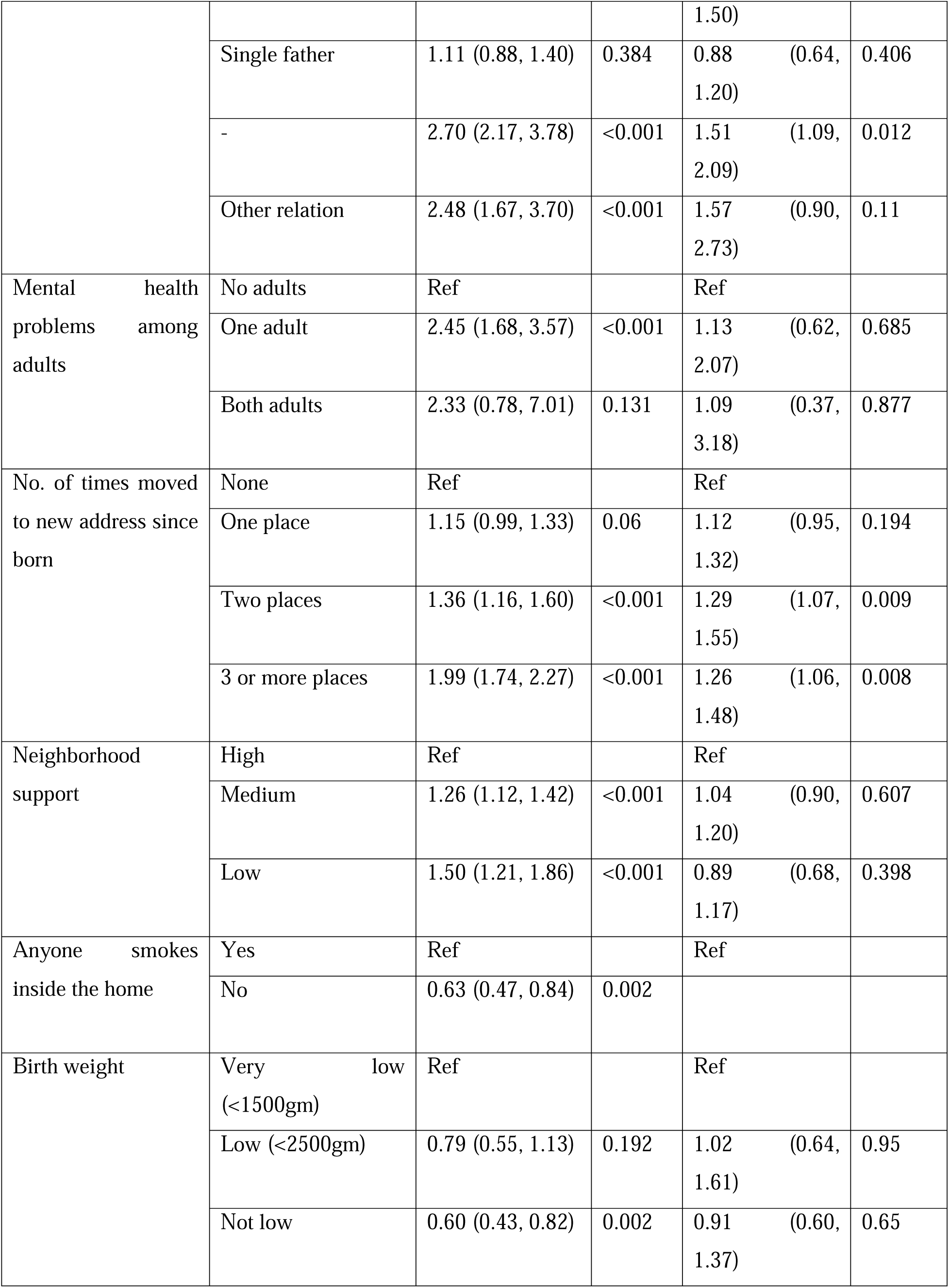
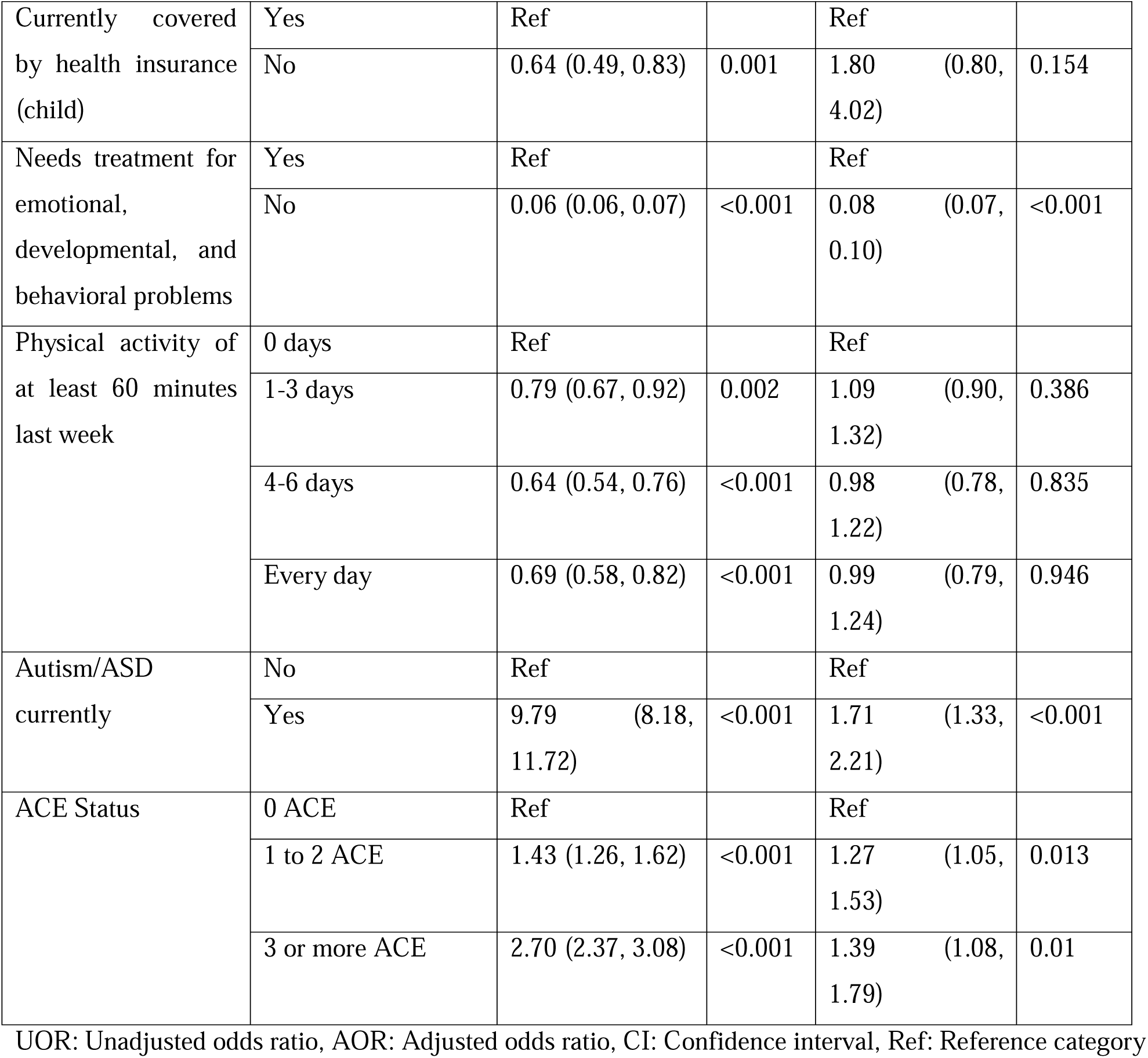
Unadjusted and adjusted associates of ADHD/ADD among 6–17-year-old U.S. children, pooled estimates of NSCH 2020-2021, N=60,809.

Additionally, the children who live in grandparent households had a 1.51 (95% CI: 1.09, 2.09, p=0.012) times higher probability of having ADHD/ADD than those who live with two biological/adoptive parents presently married.

The children who had moved to two new places since birth had a 1.29 (95% CI= 1.07, 1.55) higher likelihood of having ADHD/ADD. Moreover, the children who had moved to three or more new addresses since birth were 1.26 (95% CI: 1.06, 1.48) times more likely to experience ADHD/ADD than those without a history of changing addresses since birth. Additionally, children diagnosed with Autism/ASD at the time of data collection had a 1.71 (95% CI: 1.33, 2.21) times higher probability of having ADHD/ADD. Finally, the odds of experiencing ADHD/ADD were 1.269 (95% CI: 1.05, 1.53) times higher among children with one to two Adverse Childhood Experiences (ACEs) than those without ACEs. Likewise, children with a history of three or more ACEs had a 1.39 (95% CI= 1.08, 1.79) times higher likelihood to experience ADHD/ADD than children without any ACEs history.

## Discussion

This research examined the multifaceted relationship between determinants such as age, income level, environment, and mental well-being in children aged 6 to 17 currently residing in the United States. The study revealed significant connections between these factors and depression, anxiety, behavioral issues, and ADHD/ADD. These findings delve into understanding the influences of these determinants on children’s health and can inform intervention and policy-making strategies.

### Sociodemographic Factors and Child Mental Health

This study highlighted gender as one of the major determinants of mental health consequences. The findings suggested that the odds of depression and anxiety are higher among female children than male children, which aligns with existing literature emphasizing gender inequalities and disparities in mental health among adolescents ^6,28,29^. Cyranowski et al. (2000) mentioned that by the time they reach 15 years old, female children are about two times more inclined to encounter a depressive episode than their male counterparts, and this disparity continues for around the next three to four decades of their lives ^28^. They also highlighted that this surge failed to mirror an equal magnitude in adolescent males. McLean et al. (2011) found that the likelihood of experiencing anxiety disorder is more common among females across various age groups ^29^. In addition, Merikangas et al. (2010) also reported that all anxiety disorder subtypes were more frequently observed in females ^6^. Besides, our study also determined that age significantly influences children’s mental health. According to the study findings, adolescents aged 12-17 were more likely to experience depression and anxiety than children aged 6-11. This age-related disparity underscores the latent effects of developmental changes, school-related stressors, and peer relationships on adolescent mental health. A nationally representative prevalence study on U.S. adolescents acknowledged adolescence as the most susceptible period to developing depression and anxiety disorders^6^. The findings reported that the rate of mental disorders accelerates with age, almost doubling between the age group of 13-14 years to 17-18 years age groups. These findings are pivotal for tailored interventions focusing on age and gender, specifically focusing on age-specific challenges of adolescent girls to foster better mental health pathways.

### Ethnic/ Racial Disparities and Child Mental Health

The study highlighted disparities in mental health among ethnic and racial groups. The likelihood of experiencing depression and anxiety was lower among non-Hispanic Asian and Hispanic/Latino children than non-Hispanic White children. Additionally, it is noticeable that Black or African-American children have lower odds of facing anxiety and ADHD/ADD. Lower rates of mental health disorders among minority children may be because the outcome variables in this study were determined by yes/no questions (Has a doctor or other health care provider told you that this child has Depression/Anxiety/ADHD/Behavior problems currently). This may classify many children with the outcomes as healthy who didn’t go to a doctor or healthcare provider, particularly racial/ethnic minority children and children with lower socioeconomic status. There is loads of evidence that there are significant disparities in accessing mental health treatment in the U.S. by race/ethnicity and socioeconomic status ^12,13^.

The level of stigma surrounding health can vary among communities, especially in minority populations, which in turn affects how people seek care for their mental well-being. Stevens Watkins and colleagues (2014) highlighted the stigma associated with health in Black or African American communities ^30^. This stigma may result in individuals reporting their struggles or seeking help potentially skewing the prevalence data for this minority population. Moreover, certain cultural and societal perspectives on health might discourage the minority population from seeking assistance. Limited access to quality healthcare can also contribute to underdiagnosis within minority communities ^30^. Additionally, Sirin and colleagues (2015) argue that when minorities seek care, they may encounter professionals who lack training in approaches, leading to misdiagnoses or inadequate treatment ^31^. Cultural values such as respect, strong family bonds, interdependence, and community support play roles in safeguarding against potential mental health issues. Within communities, specifically resilience and shared support from the community can help mitigate the impact of ongoing psychosocial challenges on mental well-being, especially in environments with frequent psychosocial stressors. Therefore, it is crucial to recognize contextual factors’ influence in shaping children’s mental health outcomes. To address this issue, Lindsey et al. (2010) advocate for cultural competence training for health professionals to improve diagnostic accuracy and treatment outcomes ^32^.

Another view is race, ethnicity, and culture might act as protective factors within these ethnic communities in shaping children’s mental well-being. Lindsey et al. (2010) underscored the pivotal and protective effects of family social support in dwindling the depressive symptoms among African American adolescent boys, especially when they experienced mental health stigma ^32^. However, due to the intersectionality of their gender and racial identities, African-American women are more susceptible to psychological distress ^30^.

Integrating these elements into strategies focused on comprehending and mitigating disparities in child mental health issues is crucial. The key lies in employing tailored culturally sensitive interventions and considering diverse knowledge, experiences, and beliefs. Building partnerships among healthcare providers, educators, and community leaders is vital for establishing healthcare frameworks that are responsive to individual needs.

### Neighborhood Support, Physical Activity, and Child Mental Health

Along with the sociodemographic and racial factors, a supportive community environment appeared to be one of the most significant determinants of mental health outcomes. The study findings suggested that the higher the levels of neighborhood support, the lower the odds of anxiety and behavioral problems, implying that neighborhood support can safeguard against mental health challenges. Similar findings were reported by several studies done in previous years, where an inverse relationship was established between neighborhood support and child emotional disorders ^20,33–35^, however, these associations are significant and robust among the children of the 12 to 17-year age group ^20^. These studies demonstrated that community support and children’s engagement in the community can act as a protective factor by mitigating childhood mental ailments ^33,35^.

Our study also highlighted the positive reverse correlation between physical activity and the odds of depression and anxiety disorders, inferring that the more physically active the child is, the lower the odds of experiencing depression and anxiety become. This finding underscores the entwined connectedness of children’s physical and mental health. According to a meta-analysis, engaging in activity can result in a moderate reduction in depression and a decrease in anxiety ^36^. Therefore, encouraging activity within schools and local communities could be a successful approach to confronting mental health issues. Therefore, integrating neighborhood support elements and physical activity into approaches should be pivotal in promoting children’s well-being.

### Adverse Childhood Experiences (ACEs) and Child Mental Health

This study found significant and consistent associations between Adverse Childhood Experiences (ACEs) and several child mental health ailments. Children who experience more ACEs are more likely to show signs of depression, anxiety, behavioral issues, and ADHD/ADD. These findings are consistent with the previous literature on the long-term effects of adversity in childhood on children’s mental health and well-being. Hughes et al. (2017) reported that children with four ACEs are more at risk of various health issues than those without ACEs ^37^. In their study, Khanijahani and Sualp (2022) highlighted that children without ACEs experience little or no mental disorders. They also mentioned that the particular types of ACEs impact the mental disorders found in children. Interestingly, it was found that children who have experienced three or more ACEs are most likely to have mental health issues, with 39.4% being affected^20^. Therefore, building support networks within families, improving parenting abilities, and incorporating trauma-informed approaches in schools and healthcare settings are crucial in reducing the lasting effects of childhood adversity.

### Implications and Future Directions

The findings of this study have several implications for policy, practice, and future research. This study emphasizes the importance of creating policies and initiatives that support the neighborhood and environment in which child development occurs. Therefore, implementing socially and culturally sensitive interventions to encourage neighborhood support and physical activity to enhance child mental health promotion is crucial. Improving safety measures and recreational facilities can encourage a sense of community connection. Launching public awareness campaigns to reduce mental health stigma and educate parents, caregivers, and educators on recognizing and seeking help for mental health conditions is crucial ^20,38^. Special attention should be given to children with several ACEs who are susceptible to serious mental health illnesses. Addressing socioeconomic disparities, implementing school-based interventions like social-emotional learning programs, and fostering community-based support initiatives are vital steps. Integrated healthcare approaches, research advocacy, and policy measures can further strengthen child mental health support. Encouraging parent-teacher collaboration, reducing environmental stressors, and promoting peer support programs can contribute to holistic care.

### Strengths and Limitations

The study leverages the nationally representative and extensive National Survey of Children’s Health (NSCH) dataset, facilitating a comprehensive analysis of child mental health in the United States. It takes a holistic approach, considering various factors like demographics, socioeconomics, family, and health variables, offering a thorough understanding of the intricate factors shaping children’s mental well-being. The research findings have the potential to inform evidence-based policies and interventions aimed at improving child mental health in the United States, making a positive impact on the well-being of children. However, this study has several significant limitations that we have to consider carefully. First, the outcome variables were determined by yes/no questions (Has a doctor or other health care provider told you that this child has Depression/Anxiety/ADHD/Behavior problems currently). This may classify many children with the outcomes as healthy who didn’t go to a doctor or healthcare provider, particularly racial/ethnic minority children and children with lower socioeconomic status. Second, the study heavily relies on self-reported data, which may be subject to recall bias or underreporting of sensitive information, particularly when it comes to mental health issues. Third, the cross-sectional design of this study limits our ability to establish any causal relationships between all variables and mental health outcomes in children.

## Conclusion

This study sheds light on critical factors influencing the mental well-being of children aged 6 to 17 in the United States. Gender and age play pivotal roles, with girls and adolescents facing higher risks of depression and anxiety. Neighborhood support and physical activity are protective factors, emphasizing the importance of community engagement and active lifestyles. Adverse Childhood Experiences (ACEs) have lasting effects on child mental health, highlighting the need for early interventions and support networks. These findings have substantial implications for policy and practice. Policymakers should prioritize creating supportive environments for child development, addressing disparities, reducing stigma, and promoting culturally sensitive interventions. Collaboration among healthcare providers, educators, and community leaders is key to improving children’s mental health. Future research should explore causal relationships, delve into the intersectionality of gender and race, and investigate the long-term effects of ACEs. Longitudinal studies can provide valuable insights into how mental health conditions evolve in children over time, guiding effective interventions and policies.

## Data Availability

The dataset is publicly available and cited in the manuscript. The subset of the dataset that this study specifically analyzed can be shared upon reasonable request.

https://www.census.gov/programs-surveys/nsch.html

## Author Contributions

MM, SM, SN, and RM contributed to the study’s conceptualization, literature review, document review, development of the analysis plan, and manuscript drafting. EU, YFP, and TM provided supervision throughout the project, including guidance on the study design, analytic approach, and writing. All authors contributed to critical manuscript revisions and approved the final version for publication.

Ethical Approval: The project details were submitted to the Institutional Review Board of the University of Texas at El Paso, and they approved the study.

